# Cognitive and Motor Impairment Patterns among HIV+ Adults with and without Stroke in Botswana: Implications for Neurorehabilitation

**DOI:** 10.64898/2026.10.07.26364959

**Authors:** Michelle J. Johnson, Rochelle Mendonca, Frances S. Shofer, Ntsatsi Mogorosi, Ngadzi Goitsemang, Lefika Bothomilwe, Cassandra Ocampo, Billy Tsima, Justus Mackenzie Nthitu, Lingani Mbakile-Mahlanza, Maikutlo Kebaetse

## Abstract

**Background:** HIV and stroke are major contributors to disability in sub-Saharan Africa, yet their combined impact on cognitive and motor functioning remains poorly understood. Botswana has a high prevalence of HIV and rising stroke incidence, underscoring the need to characterize impairment to inform neurorehabilitation.

**Objective:** To examine differences in cognitive performance, motor function, and activities of daily living between adults living with HIV only and those living with both HIV and stroke in Botswana.

**Methods:** Thirty-three adults (19 HIV only; 14 HIV/ stroke) completed standardized cognitive (e.g. MoCA and Digit Symbol Coding [DSC]), and motor assessments (such as Box and Block Test [BBT], Grooved Pegboard [GPB], Grip Strength [GS], 10-Meter Walk Test [10MWT], Timed Up and Go [TUG]), and ADL measures (such as Barthel Index, Lawton IADL). Group comparisons used Wilcoxon rank-sum tests. Impairment was determined using both international z-score norms and Botswana-specific cutoffs from a prior cohort study.

**Results:** Cognitive impairment was evident in both groups, but DSC z-scores were worse in the HIV/stroke vs HIV only group (median= -2.71 vs -1.34, p=0.015). Gross motor function showed deficits in the HIV/stroke group, including lower dominant-hand BBT performance (median z-score −2.79 vs. −1.75; p=0.008). Fine motor dexterity was markedly reduced in the HIV/stroke group, with GPB dominant-hand median z-scores of −4.26 vs. −1.40 (p=0.038) and non-dominant-hand impairment rates of 100% vs. 63.2% (p=0.043). Non-dominant GS was weaker in the HIV/stroke group (100% impaired vs. 47.4%; p=0.0014). Lower-limb performance was substantially worse, including TUG (median z-score −3.63 vs. 0.10; p=0.0001) and 10MWT (median z-score −5.36 vs. −2.47; p=0.0025). ADL scores were also lower in the HIV/stroke group compared to both Barthel and Lawton (p=0.002).

**Conclusions:** HIV alone is associated with cognitive and motor impairment, with the addition of stroke producing significantly greater deficits across all motor domains. These findings demonstrate the need to integrate structured neurorehabilitation into HIV clinical care in Botswana.

## 1.0 Introduction

Human immunodeficiency virus (HIV) is an infection that attacks the immune system and weakens a person’s defenses against opportunistic infections.^1,2^ With an estimated 39 million adults and children living with HIV infection, the HIV/AIDS epidemic remains a global public health concern.^1^ Two thirds of people living with HIV (PLHIV) are in the World Health Organization African Region, particularly in sub-Saharan Africa.^1–3^ Botswana, which is located in this region, has the third highest HIV prevalence globally—about 20.5% of its population in 2022, representing approximately 320,000 people.^4^

Neurological complications of HIV, collectively termed HIV-associated neurocognitive disease (HAND), occur in 30–50% of PLHIV and limit daily activities and quality of life^5–9^ HAND affects cognition, gait, and fine motor hand function and although ART can reduce its presence, it rarely eliminates it entirely. Lawler and colleagues^10–11^ found that 37% of ART-treated PLHIV in Botswana met criteria for cognitive impairment, with significant deficits in executive function, language, memory, fine motor function, and psychomotor speed, which is consistent with neurocognitive impairment rates of 15–69% reported among ART users in other low-to-middle-income countries^5,12,13^.

ART has transformed HIV into a manageable chronic condition, shifting morbidity toward cardiovascular and cerebrovascular complications^4–6^. HIV elevates stroke risk through opportunistic infection, vasculopathy, cardiac embolism, and coagulopathy, while combination ART further increases risk via dyslipidemia and hypertension^1–3,5,-17–22^, resulting in approximately 1.82 times higher stroke risk than HIV-negative individuals. As evidence of these trends, stroke incidence is increasing among PLHIV in several sub-Saharan African countries including Botswana, Malawi and South Africa^24–26^. PLHIV and stroke in African countries are also younger than their counterparts in high income countries, such as the USA, and were less likely to have common risk factors, such as hypertension as those with stroke and no HIV.^21–24^

Bearden and colleagues^24^ investigated the association between HIV and an increased risk of stroke among adults in Botswana and reported that HIV+ adults were more likely to have a stroke at a younger age and 67% had small vessel lacunar strokes compared to HIV-negative stroke patients. While HIV reduced a stroke survivor’s overall function on activities of daily living, in Botswana, both groups were typically discharged without inpatient rehabilitation and had limited access to outpatient therapy. Hanass-Hancock and Carpenter^26^ reported after a secondary analysis of the Global Burden of Disease 2017 data, exploring the change over time in Disability Adjusted Life Years per 100 000 people over the period 1990–2016, that similar to countries in Sub-Saharan Africa and unlike Global trends, in Botswana, the main causes of years of life lost and increases in years lived with disabilities were due to HIV, cardiovascular diseases, and injuries.

The upper extremity plays an important role in enabling independent living, yet little is known about the combined impact of HIV and stroke on upper extremity function^27–29^. This study examined a Botswana-based cohort of PLHIV with and without stroke, extending Lawler’s neurocognitive battery to include upper extremity strength and gross motor measures, to explore whether stroke further compounded motor and cognitive impairments beyond HIV alone. Specifically, we explored: whether: 1) Participants living with HIV and stroke would perform worse on motor and cognitive function tests compared to those living with HIV only and 2) Participants living with HIV only or living with HIV and stroke would have similar motor and cognitive impairment trends as reported by Lawler and colleagues.^11^

## 2.0 Methods

### 2.1 Study Site

The study was carried out in a rehabilitation laboratory located at Sir Ketumile Masire Teaching Hospital (SKMTH) in Gaborone, Botswana and in the Occupational Therapy Department of the Mahalapye District Hospital. The two sites are 200 km apart, with each site drawing participants from a radius of 50-100 km.

### 2.2 Participants

A convenience sample of participants who were 18 years or older, had HIV+, were on ART for at least 6 months, medically stable and able to withstand 2-3 hours of exercise per day were recruited and enrolled between January and December 2023. Participants who had a diagnosis of stroke were included if the diagnosis of stroke was older than three months. Participants were excluded if they had severe depressive symptoms on Beck’s depression inventory (BDI) scale^31^. Potential study participants were phone-screened and if eligible were invited into the study. All participants gave informed consent in either English or Setswana according to their preference. The Institutional Review Boards of the University of Botswana (#UBR/RES/B10/032), the University of Pennsylvania (#821195), and Botswana’s Ministry of Health and Wellness (#HPDME 6/14/11) approved the study.

### 2.3 Cognitive and Motor Assessment

Participants underwent one clinical and one robot assessment within 11 days of each other. Cognitive deficits were assessed using a neurocognitive battery similar to the one used by Lawler and colleagues^11^ and augmented with an additional overall assessment of cognition using the Montreal Cognitive Assessment tool (MoCA). The cognitive battery chosen evaluated language, executive function, processing speed, attention, working memory, visuospatial functioning, orientation, and information processing, Tests included: 1) Digit Symbol Coding (DSC) ^33^, 2) Animal Fluency^34,35^, 3) Color Trails 1 & 2 (CTT1 and CTT2)^36^, and 4) the International HIV Dementia Scale (IHDS)^37^. All evaluations were administered in English or Setswana by trained assessors.

Upper limb motor function assessments were similar to those used by Bui and colleagues^30^ to assess gross motor function and strength, and additional test of upper and lower limb motor function often used in stroke. These included: 1) Box and Block Test^38^ (BBT), 2) Grooved Pegboard^39^ (GPB), 3) Grip Strength^40^, 4) 10 meter walk test^41^ (10MWT), and 5) Timed Up and Go^42^ (TUG). Participants with HIV/stroke were also evaluated with the Upper Extremity Fugl-Meyer^43^ scale and the Ashworth Scale^44^. For timed assessment, such as CTT1, CTT2, and GPB, a maximum cutoff time was used; these times were 350 seconds for CTT1 & 2 and 300 seconds for GPB. Lastly, participants were asked to report on quality of life and daily life function using the Barthel Index^45^ and IADL-Lawton-Brody^46,47^.

### 2.4 Data Analysis

Descriptive summary statistics were used to characterize the study population, using mean ± standard deviation or median and interquartile range for continuous variables, such as age, raw motor or cognitive scores, and frequencies and percentages for categorical variables, such as sex, education, or language. We determined whether participants were impaired using two methods. In method 1, where relevant, raw cognitive and motor scores were normalized using one or more demographic modifiers such as age, sex, and education, and z-scores were calculated from reported tables outside of Botswana.^48–53^ Mild impairment was identified as participants with z-scores -1.0 to -2.0, moderate -2.0 to -2.9 and severe ≤ -3.0^21^. In method 2, we used raw data cutoff data for impairment as reported by Lawler^11^ from the HIV-controls evaluated in Botswana for similar tests. We then compared the results of the two methods for tests that were similar. To compare HIV+ to HIV+/stroke participants on the cognitive and motor z-scores, Wilcoxon rank sum tests were used since values were not normally distributed. To compare the percentage impaired to the Lawler study^11^, Fisher’s exact tests were used. To assess the relationship between the Lawton and Barthel ADL scales and the motor and cognitive raw scores, linear correlations were evaluated, and multiple regression models were developed. All analyses were performed using SAS statistical software (version 9.4, SAS Institute, Cary NC). Figures were generated using GraphPad Prism (version 10.4.1, GraphPad Software LLC, San Diego, CA, USA).

## 3.0 Results

### 3.1 Participant Demographics

The study enrolled 33 HIV positive participants: 19 with only HIV (57.5%) and 14 with both HIV and stroke (42.4%) (Table 1). The average age was 49.5 years, with 60.6% females and 51.5% employed. Significant gender differences existed between groups (78.5% versus 35.7%; p=0.03), but ages were similar (p=0.16). Most had at least primary education, with higher education levels in the HIV-stroke group. Depression levels were generally normal-to-mildly depressed, without significant cohort differences. Overall cognitive impairment (MoCA score: 20.3) was moderate and similar across groups. HIV Dementia scores were lower in the HIV+/stroke group (p=0.015), which overall had moderate upper limb function impairment (UE Fugl-Meyer score: 40.3) and mild to no spasticity.

**Table 1:**
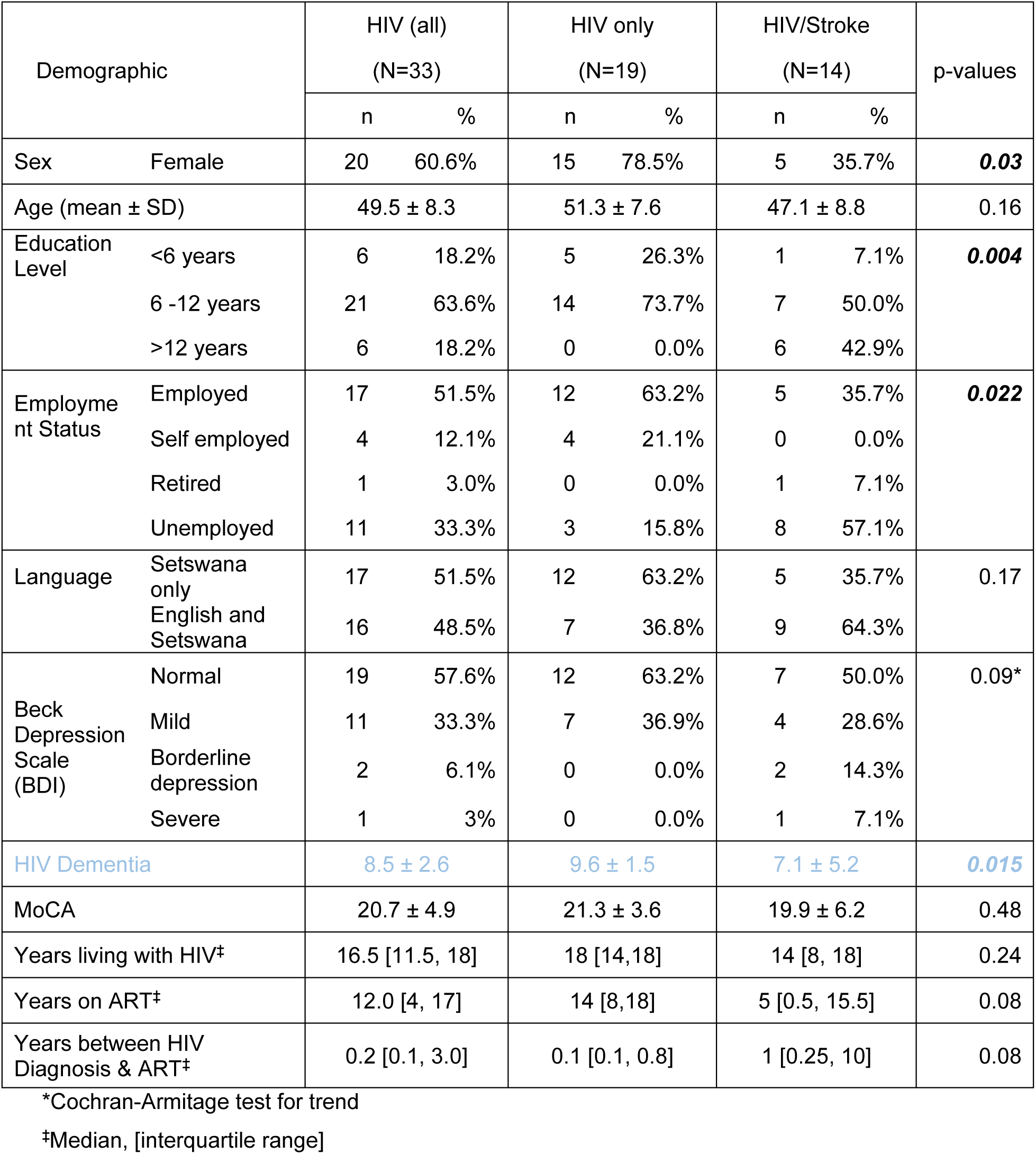
Demographic and clinical characteristics of study participants Data are presented as n (%) for categorical variables, mean ± standard deviation for normally distributed continuous variables, and median [interquartile range] for non-normally distributed continuous variables. Statistical comparisons between HIV only and HIV/Stroke groups were performed using chi-square tests for categorical variables, independent t-tests for normally distributed continuous variables, and Mann-Whitney U tests for non-normally distributed variables. *Fisher’s exact test used due to small, expected cell counts. ‡Non-parametric data presented as median [IQR]. ***Statistically significant at p < 0.05. Significant differences were observed in sex distribution, education level, employment status, and HIV Dementia Scale scores between groups.

Median number of years living with HIV was 16.5 years and on ART for 12 years. Seventy percent started ART within one year of diagnosis. ART regimens included Tenofovir combinations (60% or 20/33) and Truvada (15.1%). Five participants (15%) had a history of alcohol/drug use and/or history of tobacco use.

### 3.2 Impairment determination using normalizations tables outside of Botswana

Both HIV only and HIV/Stroke participants had impairment in cognitive domains ranging from normal (z>-1.0) to severely impaired (z < -3.0, Figure 1). Participants with HIV/Stroke performed worse on all cognitive tests compared to those with HIV only; however, only their performance on the information processing test DSC was significantly different (median difference=1.08, p=0.015). The percentage of HIV only and HIV/Stroke participants, together with cognitive impairments below a z-score of - 1.0, ranged from 52% - 79%. However, there were no significant differences between the 2 groups (Table 2). For upper limb motor scores (BBT, GPB, GS), HIV/stroke participants performed significantly worse compared to HIV only participants (Figures 2A–2C) in the non-dominant hand. Similarly, for the dominant hand, fine manipulation scores (BBT & GPB) were also worse in the HIV/stroke group, but grip strength was similar (Figure 2B). The hypothesis that those who had HIV+&Stroke would perform significantly worse than those who had HIV+ only on upper limb motor tasks was confirmed for gross manipulation tasks with both limbs and fine manipulation tasks with both limbs and grip strength with the non-dominant limb.

**Figure 1:**
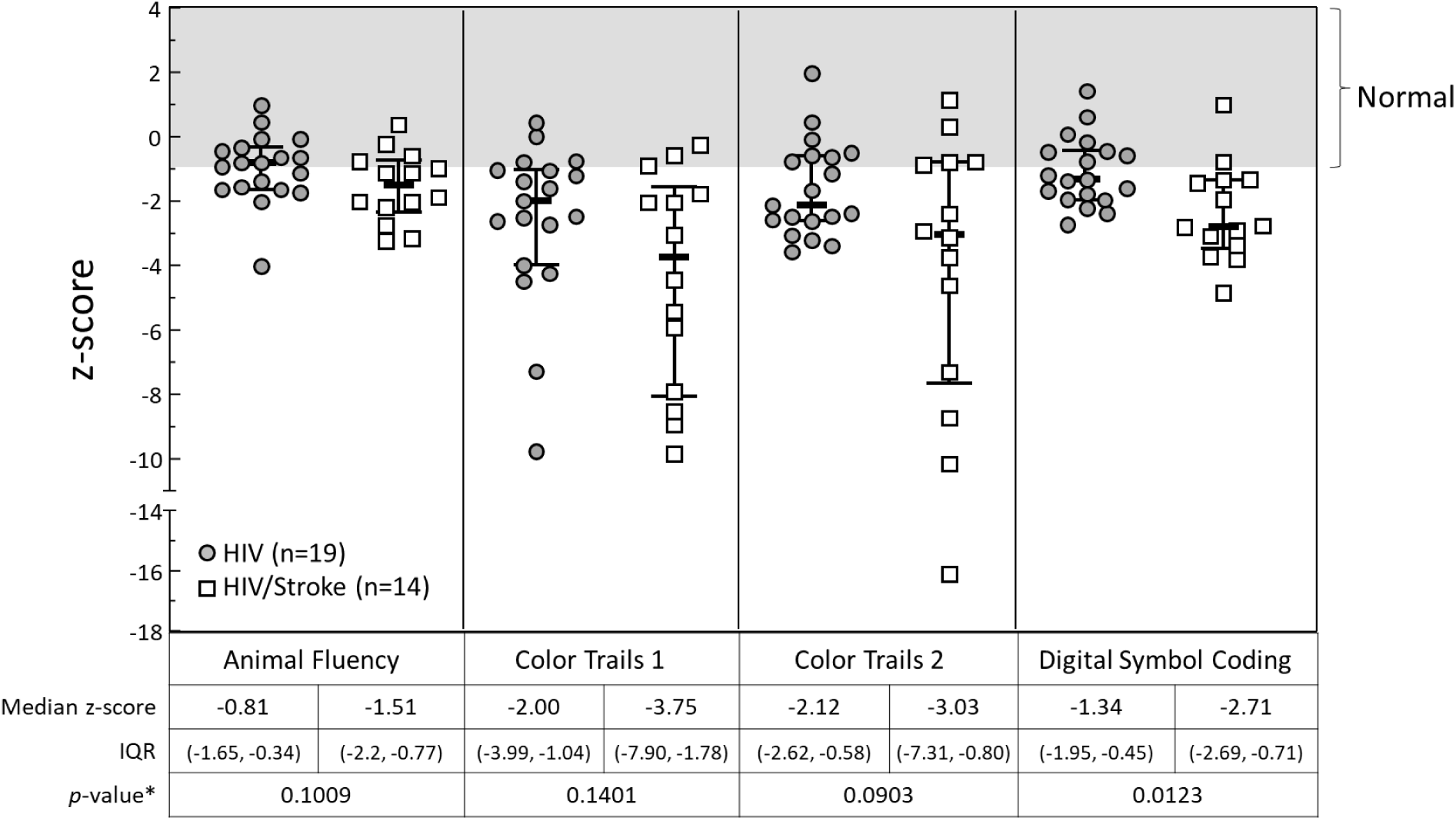
Standardized cognitive performance scores (z-scores) across four neuropsychological assessments comparing HIV only and HIV/Stroke groups. Individual data points are shown with group medians and interquartile ranges for the HIV only group (circles: n=19) and the HIV/Stroke group (squares: n=14). The shaded area indicates the normal range (z-scores > -1). Both groups performed below normal on most measures, with the HIV/Stroke group showing consistently lower median scores across all tests.

**Figure 2:**
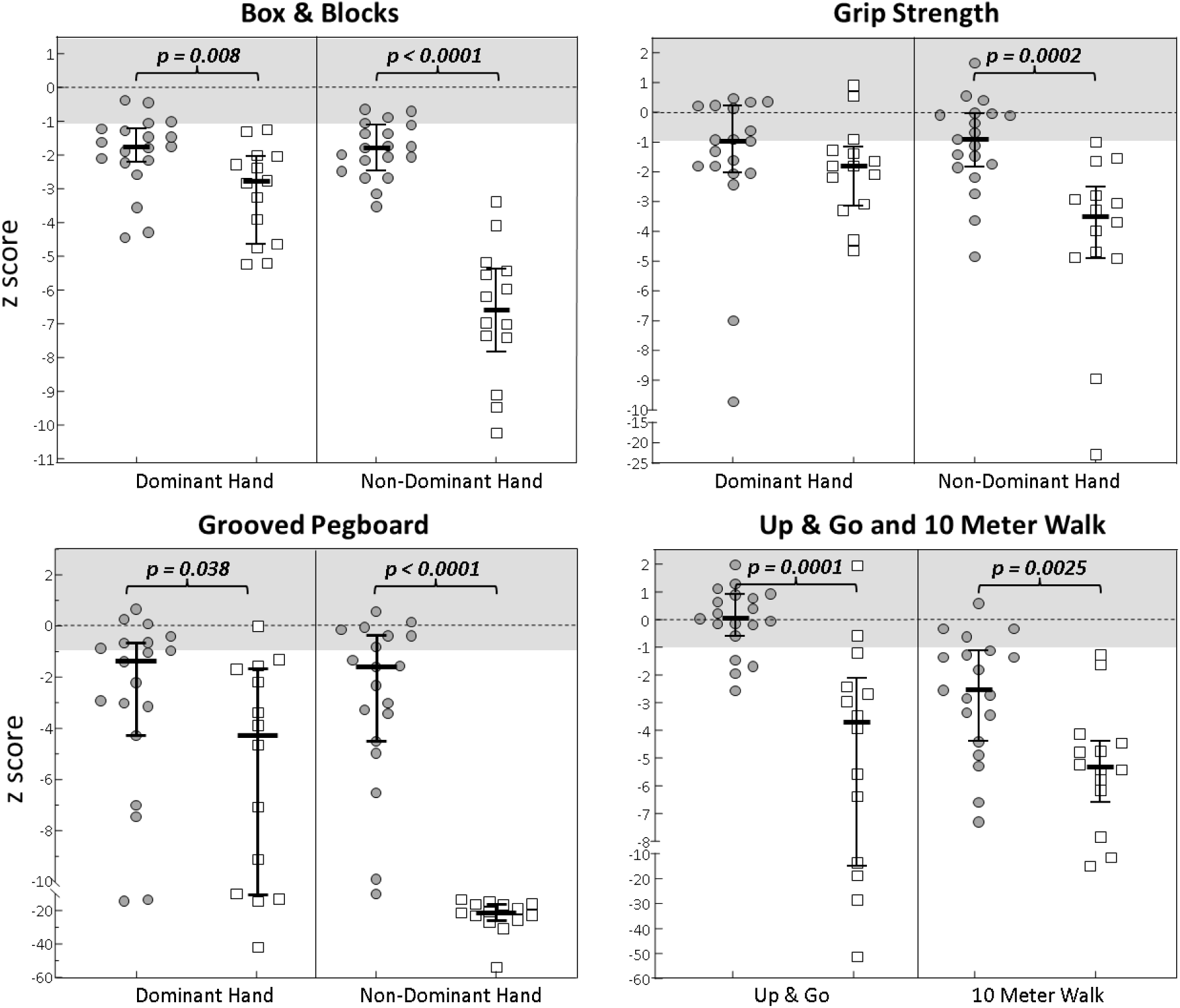
Standardized performance scores (z-scores) across motor and mobility assessments comparing HIV and HIV/Stroke groups. Top row: A. Box & Blocks and B. Grip Strength. Bottom Row: C. Grooved Pegboard and D. TUG and 10meter Walk. The individual data points are presented as normalized z-scores for HIV only (circles) and HIV/Stroke groups (squares) and are shown with group median score and interquartile ranges. The gray band highlights subjects who were considered normal in function for a particular each measure. The HIV/Stroke group demonstrated significantly impaired performance compared to the HIV only group across all measures. The mild to severely impaired participants have z-scores less than -1.0.

**Table 2:** Percentage below normal based on standardized data outside Botswana.

| Frequency & Percent below Normal<br>(z-score < -1.0) | HIV+ (All)<br>(n=33) |  | HIV only<br>(n=19) |  | HIV/Stroke<br>(n=14) |  | p-value |
| --- | --- | --- | --- | --- | --- | --- | --- |
|  | n | % | N | % | n | % |  |
| **Cognitive** |  |  |  |  |  |  |  |
| Animal Fluency: Language Fluency | 17 | 51.5 | 8 | 42.1 | 9 | 64.3 | 0.2960 |
| Color Trails 1: Visual attention processing/Psychomotor speed | 26 | 78.8 | 15 | 78.9 | 11 | 78.6 | >.9999 |
| Color Trails 2: Executive Function | 21 | 63.6 | 12 | 63.2 | 9 | 64.3 | >.9999 |
| Digital Symbol Coding: Information Processing | 23 | 69.7 | 11 | 57.9 | 12 | 85.7 | 0.1312 |
| **Motor** |  |  |  |  |  |  |  |
| Box & block (BBT-DH) | 31 | 93.9 | 17 | 89.5 | 14 | 100.0 | 0.4962 |
| Box & block (BBT-ND) | 30 | 90.9 | 16 | 84.2 | 14 | 100.0 | 0.2443 |
| Grip strength dominant hand (GS-DH) | 20 | 60.6 | 9 | 47.4 | 11 | 78.6 | 0.0876 |
| Grip strength non-dominant hand (GS-ND) | 23 | 69.7 | 9 | 47.4 | 14 | 100.0 | **0.0014** |
| Grooved pegboard (GPB-DH) | 24 | 72.7 | 11 | 57.9 | 13 | 92.9 | **0.0466** |
| Grooved pegboard (GPB-ND) | 26 | 78.8 | 12 | 63.2 | 14 | 100.0 | **0.0426** |
| Timed up & go comfort speed | 16 | 48.5 | 4 | 21.1 | 12 | 85.7 | **0.0004** |
| 10M walk | 29 | 87.8 | 15 | 78.9 | 14 | 100.0 | 0.1192 |

**Figure 3:**
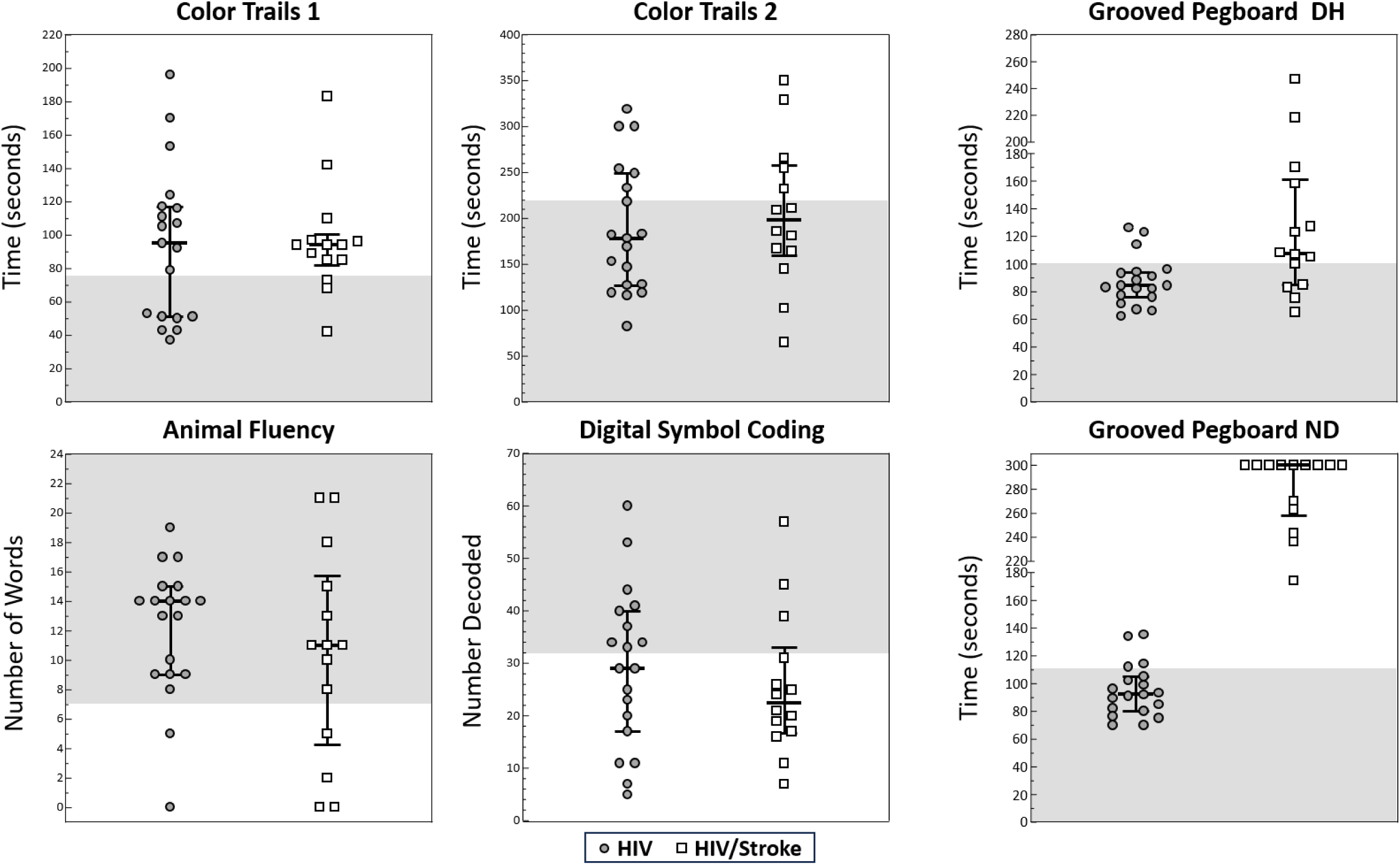
Cognitive assessment performance across six neuropsychological tests comparing HIV only and HIV/Stroke groups. Individual data points are shown with group median score and interquartile ranges. The gray band highlights the set of subjects who are considered normal in function for a particular measure based on the Lawler cutoff scores. The HIV only group (circles) consistently outperformed the HIV/Stroke group (squares) across all measures. Time-based assessments (Color Trails 1 and 2, Grooved Pegboard DH and ND) are measured in seconds, where lower scores indicate better performance. Performance-based measures (Animal Fluency, Digital Symbol Coding) are scored by number of correct responses or items decoded, where higher scores indicate better performance. DH = Dominant Hand; ND = Non-dominant/Impaired Hand.

For lower limb motor scores (TUG and 10MWT), HIV/stroke participants performed significantly worse compared to those with HIV+ only (Figures 2D). The percentage of HIV+ only and HIV/Stroke participants considered below normal was significantly different only for TUG (HIV+ only: 21.1% HIV/Stroke: 85.7%, p=0.0004). Participants with HIV/Stroke tended to have significantly lower function in their walk speed and balance scores compared to those with HIV+ only (10WMT: p=0.016 and TUG: p=0.009). The hypothesis that those who had HIV/Stroke would perform significantly worse than those who had only HIV+ was confirmed for these lower limb motor measures.

### 3.3 Impairment determination using normalization values from Botswana

Lawler and colleagues^11^ used a standardized neuropsychological test battery to explore the prevalence and features of HIV-associated neurocognitive disorders (HAND) in Botswana (see Table 2). They evaluated 60 HIV+ individuals on ART (Age: 37.5±6.2; Education years: 8.6±4.2) and compared their performance to demographically matched HIV-seronegative controls (Age: 35.8±6.7; Education years: 9.3±4.2). The average time since diagnosis was 4.6±2.5 with about 4 years on ART. Most participants graduated from junior school at a rate of 51.7%, while the study included people with no education level. Contrarily, in the current study, 28% of participants graduated from primary school, and 8% had no formal education. The mean age of the 33 participants with HIV only and HIV/Stroke tested here is older than those in the Lawler study. The study used the performance of the lowest 10^th^ percent of results achieved by the HIV-controls to define minimum “normal” performance and the participants scoring abnormal on 3 or more cognitive measures to designate cognitive impairment, which uses the definition used to define the presence of HAND or “asymptomatic neurocognitive impairment (ANI), which is defined as achieving a z-score of -1 or less in more than two cognitive areas when at least 5 cognitive areas are tested.

### 3.4 Impact on Instrumental and Basic ADL function

The Barthel Index and the Lawton scale assessed the impact of cognitive and motor impairment on ADL function. Both assessment tools demonstrated significantly higher scores in the HIV only group compared to the HIV/Stroke group (p = 0.002 for both comparisons) (Figure 4). The Lawton tool showed complete ceiling effects in the HIV group (median = 100, IQR 100-100), while the Barthel tool showed more score distribution in both groups (HIV only: median = 100, IQR 100-100; HIV/Stroke: median = 96.5, IQR 86-100). Among 19 participants with HIV only, 2 reported interferences with instrumental ADLs as measured by the Lawton scale, while 8 of 14 participants with HIV/Stroke reported issues. Similar trends were seen with Barthel Index with 4 of 19 HIV only participants reporting slight dependence for basic ADLs, compared to 10 of 14 in the HIV/Stroke group. The relationships between motor and cognitive impairment scores and these ADL scales varied by r-squared values from none to moderate (0.0 to 0.363), suggesting other factors, such as marital status, culture, assistive technology, and living context, could affect these outcomes. Walk speed (r^2^=0.1329, p=0.037), executive function (CCT2:r^2^=0.161, p=0.021), and the non-dominant/impaired hand’s grip strength (GS:r^2^=0.237, p=0.004) as well as fine and gross motor dexterity (GPB-ND:r^2^=0.319, p=0.006 and BBT-ND:r^2^=0.363, p=0.002) had significant relationships to Lawton ADL score. Walk speed (r^2^=0.214, p=0.0068), overall cognition (MoCA:r^2^=0.1171, p=0.051), and non-dominant hand’s fine motor skill (GPB-ND:r^2^=0.2167, p=0.0063), and the gross motor skill of both hands (BBT-DH:r^2^=0.253, p=0.0028 and BBT-ND:r^2^=0.159, p=0.0214) had significant relationships to the Barthel Index.

**Figure 4.**
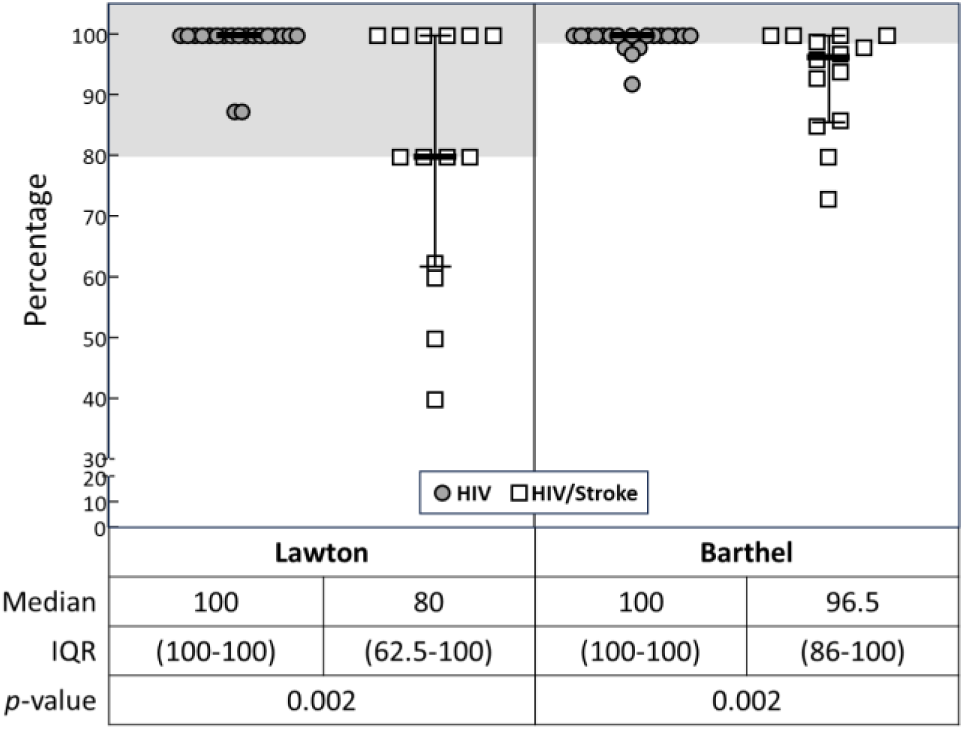
Comparison of percentage scores between Lawton and Barthel assessment tools for HIV only and HIV/Stroke patient groups. Box plots show median scores and interquartile ranges, with individual data points overlaid. The HIV only group (circles) reported lower impact on basic and instrumental ADLs than the HIV/Stroke group (squares) across all measures.

## 4.0 Discussion

Chronic complications from HIV, such as cardiovascular and cerebrovascular diseases, now account for most of the HIV-associated morbidity and mortality globally. In the USA, we examined the functional differences between a cohort of people living with HIV+ and HIV/Stroke and reported those differences in Bui and colleagues^30^, which was the first study to compare motor and cognitive function of individuals with HIV only to individuals with HIV+ and Stroke in the United States.

The study used a non-standardized neuromotor battery that examined both cognitive function and upper limb motor function. Building on work done in Bui and colleagues, the present study is one of the first studies to critically differentiate the impact of stroke on the neurocognitive and motor function in the HIV population in sub-Saharan Africa.

This paper examines the impact of HIV alone and the combined effect of HIV and Stroke on cognitive function as well as on upper and lower limb motor performance in Botswana. We uniquely identified demographic and clinical trends of a cohort of 33 people living with HIV+ (n=19 living with HIV only and n=14 living with HIV/Stroke). The age and gender trends of our cohort agreed with published literature. The HIV only cohort was on average 51.3 years old and 78.5% female, which agrees with published studies on HIV+ in sub-Saharan Africa^54^ that reports that it is not unusual for HIV+ cohorts to be predominantly female since in sub-Saharan African women are 60% more likely to contract HIV than men. Of note, the mean age of HIV/Stroke was 47.1 ± 8.8 years but only 35% female—significantly different from the HIV only—which while significantly younger than those who typically present with stroke in the USA and worldwide reflects the worldwide trend that stroke tends to affect more men than women between the age of 45 and 85 years^55^. Those with HIV/Stroke were significantly younger than their USA counterparts^30^, reflecting a trend seen before that HIV/Stroke tended to be younger and without most of the typical risk factors for stroke seen in the USA^24^. In addition, the USA cohort had significantly lower proportion of females within both sub-groups. The following sections examines the results in terms of how best to determine % impairment in the Botswana population, how stroke may impact the cognitive and motor function of an individual with HIV+, and how best to support those individuals with deficits due to HIV alone and HIV and Stroke.

### 4.1 Impairment Classification

We uniquely, quantified cognitive function and motor function of 33 people living with HIV+ (n=19 living with HIV only and n=14 living with HIV/Stroke) using a standard battery of 7 neurocognitive measures previously used with HIV+ population augmented with 4 additional upper limb measures of gross motor hand function and grip strength and 1 additional lower limb measures of balance^7–14,24,30,56,57^. Given the paucity of studies using these clinical scales in Botswana in particular and sub-Saharan Africa in general, we used two methods to characterize what constitutes normal and impaired functioning. The first method used published standardized normative data from other countries to normalize the raw scores of the data to a z-score while the second method used published raw cutoff data from a study evaluating some of the same neurocognitive measures in a HIV+ and HIV-cohort in Botswana.

Comparison with international normative data (Table 2) revealed widespread cognitive and motor impairments among HIV+ participants regardless of stroke status (n=33). Cognitive deficits were observed across all tested domains, with the highest rates in visual attention/psychomotor speed (78.8%), followed by information processing (69.7%), executive function (63.6%), and language fluency (51.5%). Motor impairments were even more pronounced, with over 90% of participants demonstrating deficits in gross motor hand dexterity bilaterally. Additional motor deficits included walking speed impairment (87.8%), fine motor dexterity deficits in both hands (72.7-78.8%), grip strength reductions (60.6-69.6%), and balance problems (48.5%).

Within this cohort, HIV+ participants without stroke comprised the majority of those with impairments in several key domains: visual attention/psychomotor speed (57.7%), executive function (57.1%), bilateral gross motor hand dexterity (dominant hand: 54.8%; non-dominant: 53%), and walking speed (51.7%). Conversely, they represented less than half of those with deficits in balance (25%), language fluency (47%), information processing (47.8%), bilateral fine motor speed (dominant hand: 45.8%; non-dominant: 46.1%), and bilateral grip strength (dominant hand: 45%; non-dominant: 39%). These findings indicate that while both cognitive and motor functioning were significantly compromised across the population, motor deficits were particularly severe, and HIV+ status alone accounted for substantial impairment burden even without concurrent stroke.

Our comparison with Botswana-specific normative data^24^ (available for 6 of 12 measures: information processing, language fluency, psychomotor speed, executive function, and bilateral fine motor skills) revealed that non-Botswana norms systematically overestimated impairment rates across all six cognitive domains (see Table 2 vs. Table 3). This overestimation was most striking for language fluency (17 vs. 6 of 33 participants classified as impaired), executive function (21 vs. 11 of 33), and fine motor dexterity in both the non-dominant hand (24 vs. 13 of 33) and dominant hand (26 vs. 15 of 33). In contrast, information processing via digital symbol coding (23 vs. 21 of 33) and psychomotor speed measured by Color Trails 1 (26 vs. 23 of 33) demonstrated relatively smaller discrepancies, suggesting these measures may be more robust to cultural and normative biases. As previously discussed, the pattern of overestimating impairment percentages persisted when analyses were restricted to the HIV+ cohort alone.

**Table 3:** Lawler vs Current study.

| Current Study Tests | Neuropsychological Measures (*by Lawler) | HIV-Impaired Cut-Off | Lawler (n=60) |  | Current Study: (n=33)<br>HIV only and HIV/Stroke |  |  | Current Study: (n=19)<br>HIV only |  |  |
| --- | --- | --- | --- | --- | --- | --- | --- | --- | --- | --- |
|  |  |  | n | % | n | % | p-value | n | % | p-value |
| Digital Symbol Coding | Information Processing Speed: Digit Symbol Coding | 32 | 19 | 32% | 21 | 58% | **0.004** | 10 | 53% | 0.111 |
| Animal Fluency | Language/Verbal Fluency: Action Fluency | 7 | 23 | 38% | 6 | 18% | 0.061 | 2 | 11% | **0.025** |
| Color Trails 1 | Visual attention and scanning/processing/<br>Psychomotor Speed: Trails A | 78.5 | 18 | 30% | 23 | 70% | **0.0004** | 12 | 63% | **0.014** |
| Color Trails 2 | Executive Function: Color Trails 2 | 221.5 | 12 | 20% | 11 | 33% | 0.209 | 6 | 32% | 0.350 |
| Grooved Pegboard DH | Fine Motor Speed: GPB Dominant Hand | 100 | 13 | 22% | 12 | 36% | 0.147 | 3 | 16% | 0.749 |
| Grooved Pegboard ND | Fine Motor Speed: GPB Non-Dom Hand | 108.5 | 15 | 25% | 18 | 54% | **0.006** | 4 | 21% | >.999 |
|  | Cognitive Impairment | >3 impaired scores | 22 | 37% | 13 | 39% | 0.826 | 4 | 21% | 0.269 |

These findings around impairment classification highlight the substantial impact of cultural and linguistic biases inherent in neuropsychological testing. Cognitive assessments developed and normed in Western populations may not adequately capture the true abilities of individuals from different cultural backgrounds, educational systems, and linguistic contexts^58^. Test materials, instructions, and cognitive strategies that are familiar and relevant in one culture may be foreign or disadvantageous in another, leading to artificially deflated performance scores. Language differences—even when tests are translated—can affect comprehension, response patterns, and cognitive processing strategies. These cultural and linguistic factors can systematically bias results, making individuals appear more impaired than they actually are when assessed against inappropriate normative standards. This underscores the critical importance of developing and utilizing population-specific normative data to ensure accurate and culturally valid assessment of cognitive function, particularly in diverse global health contexts where misclassification could have significant clinical and research implications.

### 4.2 Cognitive and Motor Impairment Trends

**Hypothesis H1** was partially confirmed since study participants who had both HIV+ and Stroke performed worse than participants with HIV only in some, but not all, assessment categories. The HIV/Stroke group tended to have similar cognitive function as their HIV only counterpart on all tested cognitive areas. ***Stroke appears to increase the burden of cognition impairment by shifting individual performances of those with HIV+ toward more severe impairment especially in the areas of language fluency, information processing and executive functioning.*** These results are supported by the stroke and HIV literature. Even after being on steady antiretroviral therapy, HIV+ is noted to create mild-to-severe neurocognitive deficits in domains such as attention, memory, psychomotor speed and executive function^59–64^. Previous investigations in both developed and resource-limited nations found that HIV infection was mainly related to executive function abnormalities.^56,62^

For example, studies in Cameroon^63^, Uganda^57^, and Nigeria^64^ show HIV infection significantly impairs executive function. Stroke compounds cognitive deficits further: in the post-acute phase, 78% of stroke patients show impairment in at least one cognitive domain, which was most commonly attention (48.5%), followed by language, memory, and executive dysfunction. At one year, attention deficits persist while executive function and memory improve^64^. A Dutch study found over 70% of stroke survivors had impaired information processing at 2 months, and nearly half still showed mild-to-moderate deficits at 5 years, primarily in processing speed and executive function^65^. Barker-Collo et al.^66^ found that 47% of stroke survivors still had mild-to-moderate impairments in information processing speed and executive function at 5 years post-stroke — consistent with Cumming et al. ^67^, who identified executive function and attention as the most impaired domains following stroke, rather than memory.

HIV causes mild-to-moderate motor deficits in walking and fine motor speed, while 40–70% of stroke survivors develop hemiparesis. As expected, the HIV/Stroke group showed significantly greater motor impairment than the HIV-only group. ***Stroke appears to increase the burden of motor impairment in those with HIV by shifting individual performances toward more severe impairment on all motor areas especially in the areas of balance, walking, and grip strength, fine motor speed and gross motor dexterity in the non-dominant hand.*** Bui et al. ^30^ reported that HIV impairs bilateral upper limb gross motor function, manifesting as difficulty reaching and manipulating large objects in those with and without stroke. Despite similar MoCA scores, the US HIV-only group showed better cognitive function and gross motor hand function than the HIV/Stroke group, though grip strength and fine motor speed were comparable. The US cohorts were older, more educated than the Botswana cohort with less cognitive impairment in information processing and psychomotor speed domains (see supplementary Tables).

**Hypothesis H2** was partially rejected. The HIV-only group in this study differed from Lawler et al.’s cohort mainly in language fluency and psychomotor speed, likely reflecting demographic differences (older age, more female, less educated). When stroke was added, differences extended to information processing and fine motor function as well.

### 4.3 Impact on ADLs

The real-world impact of the cohort’s measured functional status on basic and instrumental ADLs using self-reported clinical questionnaires were unexpected in that despite moderate to severe reports of both cognitive and motor impairments, the reported relationships with ADLs, both basic and instrumental, was milder^74–78^. The best linear regression models using cognitive and motor variables as predictors explained about 58% of the variance in basic or instrumental ADLs. Regressions, however, did show that walk speed, cognition, non-dominant grip strength, and fine and gross motor skill impacted both basic and instrumental ADLs, while gross motor skill in dominant hand impacted basic ADL’s. These results indicate that both HIV only and HIV/Stroke groups have significant problems with complex manipulation and walking tasks requiring significant cognitive as well as motor functioning. Having a stroke along with HIV appears to further impair the motor functioning in the hand and legs.

The notable disconnect between objective impairment levels and self-reported daily limitations suggests that many PLHIV may be compensating for deficits or relying on caregiver support without fully recognizing the extent of their functional decline. This underscores the importance of objective clinical assessments in identifying rehabilitation needs that patients themselves may not report. Rehabilitation programs aimed at this population should therefore target both adaptive strategies and active recovery, ensuring that functional gains are sustainable and not solely dependent on compensation.

### 4.4 Implication for Neurorehabilitation

The motor and cognitive scores highlight the need for and value of providing consistent and regular physical and cognitive rehabilitation for HIV+ only as well as HIV/Stroke. Despite prior recognition that more than 40% of PLHIV live with impairment and have difficulty with daily activities, PLHIV are not consistently prescribed neurorehabilitation. This is true in Botswana as well as in many other high, middle and low-income countries. Bearden and colleagues^24^ reported that those with stroke only as well as those with HIV/Stroke did not receive the recommended dosage of therapy, so recovery was retarded.

This study highlights a critical gap in the HIV care continuum that demands urgent attention from healthcare systems and policymakers. Given that all people living with HIV (PLHIV) demonstrated meaningful cognitive and motor impairments independent of stroke, neurorehabilitation should be reconsidered not as a specialized add-on but as a standard component of HIV care. Early and consistent integration of rehabilitation services into HIV management may help prevent functional decline and improve long-term outcomes across this population.

Finally, the relatively young age of many PLHIV presents both a challenge and an opportunity. Cognitive and motor impairments in this population carry significant economic and social consequences, including reduced workforce participation and increased dependency. Timely neurorehabilitation investment can support return to work, preserve independence, and reduce the broader societal burden of HIV-related disability — making the case not only on clinical grounds but as a sound economic and social priority.

### 4.5 Study Limitations

Given the small sample size of convenience, results should be interpreted with caution. However, we do show agreement with other studies. Additionally, the Lawton and Barthel ADL questionnaires were self-reported and are vulnerable to reporting biases. The Lawton Brody IADL scale additionally has not been updated since its first development and assumes that males do not perform certain activities such as laundry and housekeeping, which may no longer be accurate. These questionnaires were also primarily developed for a Western population and might have cultural biases regarding typical ADLs and IADLs performed by a Botswana population. Despite these limitations, these results lay the groundwork for future studies to further explore how stroke impacts cognitive and motor function of those with HIV+ and how we can develop novel methods for early detection of impairment and early and intense neurorehabilitation.

## 5.0 Conclusions

This study provides compelling evidence that comprehensive neurorehabilitation services should be integrated as a standard component of HIV care globally, particularly in sub-Saharan Africa. Chronic complications from HIV such as cerebrovascular diseases including Stroke, account for most of the HIV-associated morbidity and mortality globally, especially in low- and middle-income countries in Sub-Saharan Africa. While studies have extensively evaluated HAND in HIV+, not many have compared the impact of HIV alone and the combined effect of HIV and Stroke on cognitive function and upper and lower limb motor performance in LMICs.

Given that rehabilitation could enable workforce participation and reduce healthcare dependency, while addressing the substantial unmet needs revealed by this research, healthcare systems with significant HIV cohorts must prioritize the development and implementation of accessible, culturally appropriate neurorehabilitation programs that target the specific motor and cognitive domains most affected, recognizing neurorehabilitation as an essential component of comprehensive HIV care.

## Data Availability

De-identified data produced in the present study are available upon reasonable request to the authors

## Acknowledgements

This work was made possible through core services and support from the National Institute of Neurological Disorders and Stroke of the National Institutes of Health (T32NS091006); the Botswana UPenn Partnership, the Center for Global Health; the University of Pennsylvania’s Center for AIDS Research (P30AI 045008); the Fogarty International Center of the National Institutes of Health (R21TW012659); the University of Pennsylvania’s Departments of Bioengineering and Physical Medicine and Rehabilitation; Columbia University’s Program of Occupational Therapy and the University of Botswana Faculty of Medicine. We thank Sir Ketumile Masire Teaching Hospital for the provision of the laboratory space. We also thank our medical colleagues and therapy colleagues at Princess Marina Hospital in Gaborone, Mahalapye Hospital in Mahalapye, at Meribah Occupational Therapy and Stroke Rehabilitation Center and private clinics in Gaborone, which were other recruitment sites. We thank all the participants for their time and feedback. The content is solely the responsibility of the authors and does not necessarily represent the official views of the National Institutes of Health.

## Ethics Approval

The Institutional Review Boards of the University of Botswana (#UBR/RES/B10/032), the University of Pennsylvania (#821195), and Botswana’s Ministry of Health and Wellness (#HPDME 6/14/11) approved the study. Approvals were also obtained from recruitment sites. All mandatory safety procedures have been complied with in the course of conducting the work reported in this paper.

## Data Availability Statement

All data relevant to this study will be included in the article or uploaded as supplementary information in the event of publication.

